# Phase 2 Study of Sorafenib, Valproic Acid, and Sildenafil in the Treatment of Recurrent High-Grade Glioma

**DOI:** 10.1101/2024.04.23.24304634

**Authors:** Andrew S. Poklepovic, Palak Shah, Mary Beth Tombes, Ellen Shrader, Dipankar Bandyopadhyay, Xiaoyan Deng, Catherine H. Roberts, Alison A. Ryan, Daniel Hudson, Heidi Sankala, Maciej Kmieciak, Paul Dent, Mark G. Malkin

**Author notes:** **Corresponding Authors: Mark G. Malkin, MD Andrew S. Poklepovic, MD Paul Dent, PhD** School of Medicine Virginia Commonwealth University Richmond, VA, 23298.

## Abstract

Here we report the results of a single-center phase 2 clinical trial combining sorafenib tosylate, valproic acid, and sildenafil for the treatment of patients with recurrent high-grade glioma (NCT01817751). Clinical toxicities were grade 1 and grade 2, with one grade 3 toxicity for maculopapular rash (6.4%). For all evaluable patients, the median progression-free survival was 3.65 months and overall survival (OS) 10.0 months. There was promising evidence showing clinical activity and benefit. In the 33 evaluable patients, low protein levels of the chaperone GRP78 (HSPA5) was significantly associated with a better OS (p < 0.0026). A correlation between the expression of PDGFRα and OS approached significance (p < 0.0728). Five patients presently have a mean OS of 73.6 months and remain alive. This is the first therapeutic intervention glioblastoma trial to significantly associate GRP78 expression to OS. Our data suggest that the combination of sorafenib tosylate, valproic acid, and sildenafil requires additional clinical development in the recurrent glioma population.

## Introduction

High-grade gliomas are the most common malignant primary brain tumors in adults and are associated with poor prognosis [1]. Despite the administration of optimal therapy, nearly all high-grade gliomas eventually reoccur. The median survival following recurrence is only 25 to 30 weeks for World Health Organization (WHO) grade 4 gliomas and 39 to 47 weeks for WHO grade 3 gliomas [2, 3]. Glioblastomas account for approximately 60% to 70% of high-grade gliomas. Anaplastic astrocytomas comprise 10% to 15%. Anaplastic oligodendrogliomas represent 10% of high-grade gliomas. Less common tumors such as anaplastic ependymomas and anaplastic gangliogliomas make up the remainder [4].

Many targeted agents have been used to target the aberrant signaling pathways and tumor cell biology in recurrent high-grade glioma, very few of which have prolonged either progression-free survival (PFS) or overall survival (OS). Bevacizumab (Avastin®), a monoclonal antibody against VEGF-A has been studied extensively in this patient population. In a randomized, multicenter, noncomparative phase 2 trial, the estimated 6-month PFS rates for patients with recurrent glioblastoma were 42.6% for bevacizumab alone and 50.3% for bevacizumab plus irinotecan [5]. This trial did not show any OS benefit, which is the case with a range of bevacizumab and chemotherapy combinations, e.g., lomustine, carmustine, or temozolomide. However, there was an improvement of quality of life with decreased corticosteroid use [6]. Additional therapies are urgently needed for the treatment of recurrent glioblastoma.

In the US, sorafenib tosylate (Nexavar®) is approved for the treatment of differentiated thyroid carcinoma refractory to radioactive iodine treatment, and renal cell and hepatocellular carcinomas. Sorafenib is a multi-targeted protein kinase inhibitor that was originally developed as an inhibitor of RAF-1/B-RAF, components of the ERK1/2 pathway. Sorafenib was subsequently shown to inhibit multiple other kinases, including class III tyrosine kinase receptors such as platelet-derived growth factor receptors (PDGFRs), vascular endothelial growth factor receptors (VEGFRs), c-Kit, and FLT3. Our initial findings demonstrated that sorafenib played a role in the activation of death receptors resulting in tumor cell death [7]. The anti-tumor effects of sorafenib in cancer cells is synergistically enhanced by concomitant histone deacetylase (HDAC) inhibitor exposure, including the HDAC inhibitors vorinostat, sodium valproate, and entinostat [8–10]. Cell death by the drug combination is mediated through the extrinsic pathway of apoptosis, the death receptor CD95, and by macroautophagy-induced mitochondrial dysfunction.

Valproic acid (sodium valproate, Depakote®) is an anti-seizure medication and bi-polar disorder therapeutic in common medical use and is referred to as a non-enzyme-inducing antiepileptic drug (non-EIAED). It has subsequently been shown that valproic acid also acts as an HDAC inhibitor [11, 12]. Valproic acid has been demonstrated at clinically relevant concentrations to inhibit class I (HDAC 1, 2, and 3, along with HDAC 8) and class II HDACs (HDAC 4,5, 6, and 7) [11, 12]. Valproate has dose-limiting toxicity in the liver; however, it does not modulate the cytochrome p450 system; thus, valproic acid does not have many of the drug-drug interactions with standard chemotherapy agents that are common for other antiepileptic drugs.

Sildenafil (Viagra®) is FDA approved for the treatment of both erectile dysfunction and pulmonary hypertension and has been administered to men, women, and children on a chronic daily basis [13]. The primary target of sildenafil is phosphodiesterase 5 (PDE5). Inhibition of PDE5 increases the levels of its substrate cyclic GMP (cGMP) in cells resulting in the activation of protein kinase G (PKG). PKG signaling increases nitric oxide synthase (NOS) activity which induces smooth muscle relaxation resulting in vasodilation. Sorafenib and HDAC inhibitors both individually generate reactive oxygen species in tumor cells due to their effects on mitochondrial biology, which when combined with nitric oxide from sildenafil form peroxynitrite, a lethal free radical.

Sildenafil and PDE5 inhibitors may also have other functions that relate to anticancer therapy. For example, PDE5 inhibitors increase chemotherapy delivery to brain tumors in animal models, through increased tumor cGMP levels [14]. ATP-binding cassette (ABC) proteins transport various molecules across extra and intracellular membranes. Sildenafil inhibits the ABCB1 and ABCG2 drug-efflux pumps, and reverse ABCB1 and ABCG2 mediated chemotherapeutic drug resistance [15]. The ABCG2 transporter has recently been shown to be the dominant transporter that limits transport of sorafenib into the brain [16]. This is clinically relevant as glioblastoma is considered to be a diffuse disease, with many areas of tumor residing in areas of the brain behind an intact blood-brain-barrier (BBB). Inhibition of the ABCG2 transporter with sildenafil may increase sorafenib drug concentrations in the brain, improving sorafenib anti-tumor efficacy. In vitro data demonstrated that sorafenib and sildenafil synergized to kill tumor cells, which required PKG and the actions of inducible NOS [23].

Following the initiation of this trial, subsequent studies from our group demonstrated that sorafenib at in vivo physiologic nanomolar concentrations is an inhibitor of both HSP90 family and HSP70 family chaperone proteins, and in particular, the HSP70-family chaperone GRP78/BiP/HSPA5 [17, 18]. GRP78 plays a central role in sensing endoplasmic reticulum (ER) stress in cells, inhibiting the actions of PKR-like ER kinase (PERK) and inositol requiring enzyme type 1 (IRE1) [19–22]. Under conditions where denatured protein levels are high, GRP78 disassociates from PERK and IRE1 and binds to the unfolded proteins. This leads to activation of PERK and IRE1. PERK phosphorylates eIF2α serine 51, which reduces translation from ∼90% of all mRNAs, and conversely enhances translation from ∼10% mRNAs. The expression of proteins involved in protein degradation processes, e.g., Beclin1 and ATG5 in macroautophagy, is enhanced in an eIF2α-dependent fashion. In parallel, the levels of cytoprotective proteins with short half-lives such as MCL1 and BCL-XL is reduced. In vitro, the lethal interaction between sorafenib and HDAC inhibitors or PDE5 inhibitors required an ER stress signal, and over-expression of GRP78 reduced the ability of either drug combination to kill tumor cells that was associated with reduced eIF2α S51 phosphorylation and no observable changes in the protein levels of ATG5, Beclin1, MCL1, or BCL-XL.

Promising single agent in vitro activities and potentially complementary mechanisms of action suggest that the combination of sorafenib, valproic acid, and sildenafil may have therapeutic potential for the treatment of high-grade glioma in the clinic. The combination of sorafenib and valproic acid is predicated on the basis that sorafenib activity is enhanced by HDAC inhibition [9, 10, 13, 23]. The addition of sildenafil is based on its ability to increase peroxynitrate levels and to block ABCB1 and ABCG2 drug-efflux pumps increasing drug delivery to the tumor [15, 16] as the ABCG2 transporter is the primary transporter involved in the efflux of sorafenib at the BBB, blocking its action is predicted to increase the concentration of sorafenib in the brain. The study was undertaken to determine the safety profile and any survival benefit of this 3 drug combination.

## Methods and Materials

### Drug supply

Sorafenib and sildenafil commercial stock was obtained by the Virginia Commonwealth University (VCU) Massey Cancer Center and provided at no charge to study participants. The study drugs were provided through the VCU Health System Investigational Drug Service.

### Patient eligibility

Eligible patients were 18 years of age and older with an Eastern Cooperative Oncology Group (ECOG) performance status of 0-2 and pathologically confirmed high-grade glioma (WHO grade 3 or 4), with documented CT or MRI progression or recurrence and measurable or evaluable disease by Response Assessment in Neuro-Oncology (RANO) (MRI) or Macdonald (CT) criteria [24]. Additional inclusion criteria included creatinine clearance greater than 30 mL/min, aspartate aminotransferase (AST) and alanine aminotransferase (ALT) less than 3 × the upper limit of normal (ULN), and serum total bilirubin less than 1.5 × the ULN.

Patients were excluded from the study if they had any of the following: a seizure disorder necessitating the use of EIAEDs; a contraindication to antiangiogenic agents; clinically significant cardiac disease; prior allergic reaction or intolerance to components of the investigational regimen; history of priapism; systolic blood pressure (BP) greater than 160 mm Hg or diastolic pressure greater than 100 mm Hg despite optimal medical management; QTc greater than 480 ms (Common Terminology Criteria for Adverse Events [CTCAE] grade 2 or greater) on screening ECG; required treatment with strong CYP3A4 inhibitors or inducers; chronic nitrate therapy or alpha-blockers; or were pregnant or nursing.

The Virginia Commonwealth University (VCU) Institutional Review Board approved the study protocol. NCT01817751 All patients provided written informed consent.

### Study design

This open-label, single-arm phase 2 study of sorafenib, valproic acid, and sildenafil in the treatment of patients with recurrent high-grade glioma was conducted at VCU (ClinicalTrials.gov NCT01817751). The treatment schedule was a combination of sorafenib, valproic acid, and sildenafil, each agent administered orally, twice daily continuously. A cycle consisted of 4 weeks. The first cycle began once therapy began with all 3 agents—after, if necessary, a titration of the valproic acid dosage to therapeutic level.

The first 6 patients evaluable for qualifying toxicity assessment were treated as a safety lead-in. Sorafenib was dosed at 400 mg orally, twice daily, continuously. Subsequent dose modifications were allowed based on toxicity assessment. Sildenafil was dosed at 50 mg orally, twice daily, continuously. Subsequent dose modifications were allowed based on toxicity assessment. Study treatment dose modifications consisted of dose omission and/or schedule adjustment of 1 or more agents as determined clinically appropriate. In general, agent(s) were omitted pending resolution of toxicity to _≤_ grade 1 and then resumed at the same or a lower dose or frequency.

The primary objective was to determine the efficacy of the drug combination with reference to 6-month PFS. The study was conducted through an adaptive design potentially including two Simon’s two-stage mini-max designs. Initially, patients with recurrent high-grade glioma were enrolled, regardless of tumor molecular subtype, to determine the efficacy of the drug combination. If efficacy criteria were not met in patients inclusive of all molecular subtypes, then only patients with tumors that express PDGFRα would be enrolled. Therefore, two Simon’s designs would be included; the first for patients in the entire cohort and the second for patients with PDGFRα-expressing tumors only if efficacy criteria were not met in the entire cohort.

### Response and toxicity assessment

All adverse events (AEs) from the time of enrollment until the end of study were graded using the National Cancer Institute (NCI) CTCAE version 4.0. Clinical and basic laboratory assessments (liver and renal) were performed at baseline, and weekly for each cycle. Complete blood counts with differential were also examined on days 1 and 15 of each cycle.

### Immunohistochemistry: PDGFR**α** + GRP78

Tumor samples from the primary resection at the time of initial treatment for high-grade glioma were used to determine the level of PDGFRα. Anti-PDGFRα antibody D13C6 (Cell Signaling Technology) and anti-GRP78 BiP antibody ab21685 (Abcam) were used at a 1/100 dilution with the DAKO Autostainer Plus automated system. The results were scored by a pathologist as a product of the intensity (0-2 scores) and the percentage scores (with scores 0 for 0%, 1 for 1-33%, 2 for 34-66% and 3 for 67-100%). The standard definition of low (0 or 1) and high (greater than 1 to 6) was used.

### Statistical analysis

Twenty-one out of 33 evaluable patients who had non-missing PDGFRα expressions were grouped into PDGFRα-High and PDGFRα-Low based on their final PDGFRα value. The patients with a final value larger than 1 were assigned to the PDGFRα-High group, while patients with a final value 0 or 1 were assigned to the PDGFRα-Low group. The Log-rank test was used for comparing the distribution of the OS time between these two PDGFRα groups. The same grouping method was applied to assign the 28 patients who had non-missing GRP78 expressions into GRP78-High and GRP78-Low groups. The same test was used for these two GRP78 groups. All the analyses were conducted using the SAS (Statistical Analysis System, version 9.4) software. The significance level was set to 5%.

## Results

### Patient characteristics

Forty-seven patients were enrolled in the study and on treatment between July 24^th^ 2014 and April 12^th^ 2023. All 47 patients were deemed evaluable for toxicities. Out of the 47 patients enrolled in the trial, 33 patients were evaluable for objective response. Patients were treated with the combination of twice daily sorafenib and valproic acid for a minimum of 28 days and had their disease re-evaluated. The demographic characteristics of the 33 patients evaluable for objective response were comparable to the total 47 patients treated. The patients enrolled in the study were predominantly male with an ECOG performance status of 0-1. Median age at enrollment was 58. Glioblastoma was the most common histology of tumor. Of the evaluable 33 patients, 29 eventually came off treatment due to apparent disease progression, which may have been due to drug-enhanced radiation necrosis, and 3 patients came off due to toxicity (Table 1).

**Table 1:**

| Table 1 |  |  |
| --- | --- | --- |
| Characteristics | Number of Treated Patients (%)<br>(N=47) | Number of Evaluable Patients (%)<br>(N=33) |
| **Gender** |  |  |
| Female | 12 (25.53) | 6 (18.18) |
| Male | 35 (74.47) | 27 (81.82) |
| **Race** |  |  |
| American Indian or Alaska Native | 1 (2.13) | 0 (0.00) |
| Black or African American | 5 (10.64) | 4 (12.12) |
| White | 41 (87.23) | 29 (87.88) |
| **Ethnicity** |  |  |
| Hispanic or Latino | 1 (2.13) | 0 (0.00) |
| Non-Hispanic | 46 (97.87) | 33 (100) |
| **Age** |  |  |
| Mean | 58.1 | 54.85 |
| Median | 60.00 | 56.00 |
| Range | 54.00 | 51.00 |
| Min - Max | 28.00 - 82.00 | 28.00 - 79.00 |
| **Performance Status** |  |  |
| 0 | 8 (17.02) | 6 (18.18) |
| 1 | 31 (65.96) | 24 (72.73) |
| 2 | 8 (17.02) | 3 (9.09) |
| **Histology** |  |  |
| - Astrocytoma, Anaplastic | 1 (2.13) | 1 (3.03) |
| - Glioblastoma, NOS | 38 (80.85) | 26 (78.79) |
| - Gliosarcoma | 2 (4.26) | 2 (6.06) |
| - Oligodendroglioma, NOS | 2 (4.26) | 1 (3.03) |
| - Oligodendroglioma, Anaplastic | 4 (8.51) | 3 (9.09) |
| **Extent of Resection at Diagnosis** |  |  |
| Biopsy | 6 (12.77) | 3 (9.09) |
| GTR | 18 (38.30) | 12 (36.36) |
| Resection | 10 (21.28) | 7 (21.21) |
| STR | 13 (27.66) | 11 (33.33) |
| **Extent of Resection at Recurrence** |  |  |
| Biopsy | 1 (2.13) | 1 (3.03) |
| Biopsy & LITT | 2 (4.26) | 2 (6.06) |
| GTR | 3 (6.38) | 2 (6.06) |
| Resection | 5 (10.64) | 5 (15.15) |
| STR | 10 (21.28) | 6 (18.18) |
| No | 26 (55.32) | 17 (51.52) |
| **PDGF Status** |  |  |
| High | 10 (21.28) | 7 (21.21) |
| Low | 20 (42.55) | 14 (42.42) |
| Missing | 17 (36.17) | 12 (36.36) |
| **GRP78 Status** |  |  |
| High | 22 (46.81) | 17 (51.52) |
| Low | 18 (38.3) | 11 (33.33) |
| Missing | 7 (14.89) | 5 (15.15) |
| **Prior Treatment (Number of Regimens)** |  |  |
| Mean | 1.57 | 1.52 |
| Median | 1 | 1 |
| Range | 3 | 2 |
| Min - Max | 1 - 4 | 1 - 3 |

*\*GTR = Gross Total resection, STR = Subtotal Resection, Resection = GTR or STR, Biopsy = Biopsy, LITT = Laser Interstitial Thermal Therapy.*

### Survival

No difference in OS was observed comparing patients with an ECOG PS 2 vs 0 with a hazard ratio of 5.684, although the value was approaching significance (p < 0.0613). A statistical difference in OS was seen between patients with ECOG PS of 1 vs 2 with a hazard ratio of 0.141 (p < 0.0083). Thus, worse performance status was associated with worse OS outcomes.

### PDGFRα and GRP78

The expression of PDGFRα and of GRP78 were analyzed. PDGFRα and GRP78 expression was categorized as high (greater than 1 expression) or low (0 or 1 expression). OS was not significantly different between the high and low PDGFRα patient groups, although this value was also approaching significance (p < 0.0728) (Figure 1). OS was significantly different comparing high and low GRP78 expression (p < 0.0026). The hazard ratio (high vs low) was 3.819, implying that the hazard of death for patients in the GRP78 high value group was 3.819 times that for the patients in the GRP78 low value group (Figure 2).

**Figure 1:**
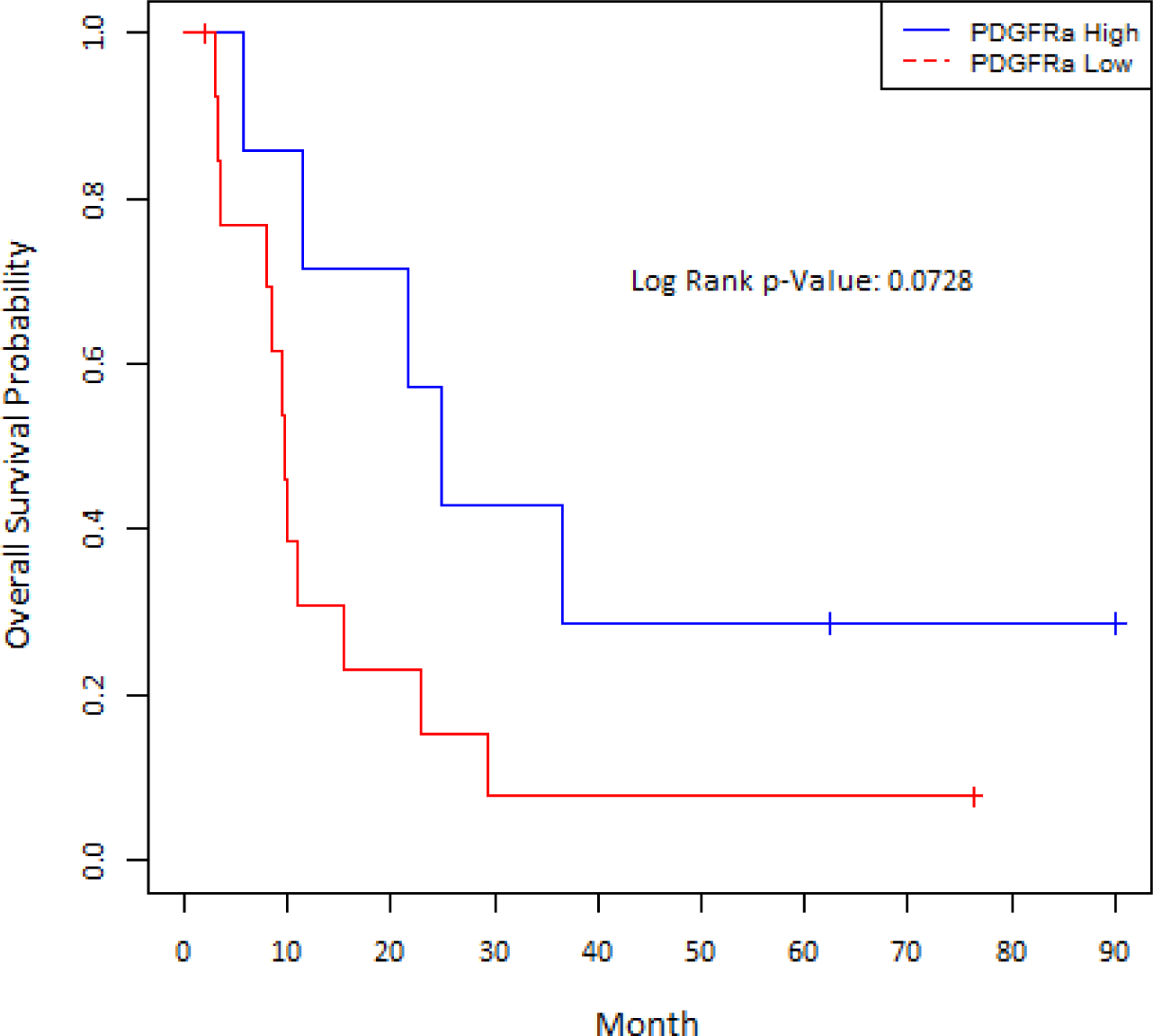
OS by PDGFRα status (high, low). P < 0.0728 of the Log-Rank Test over the strata indicates that the distributions of the OS time were not significantly different between the high and low PDGFRα patient groups.

**Figure 2:**
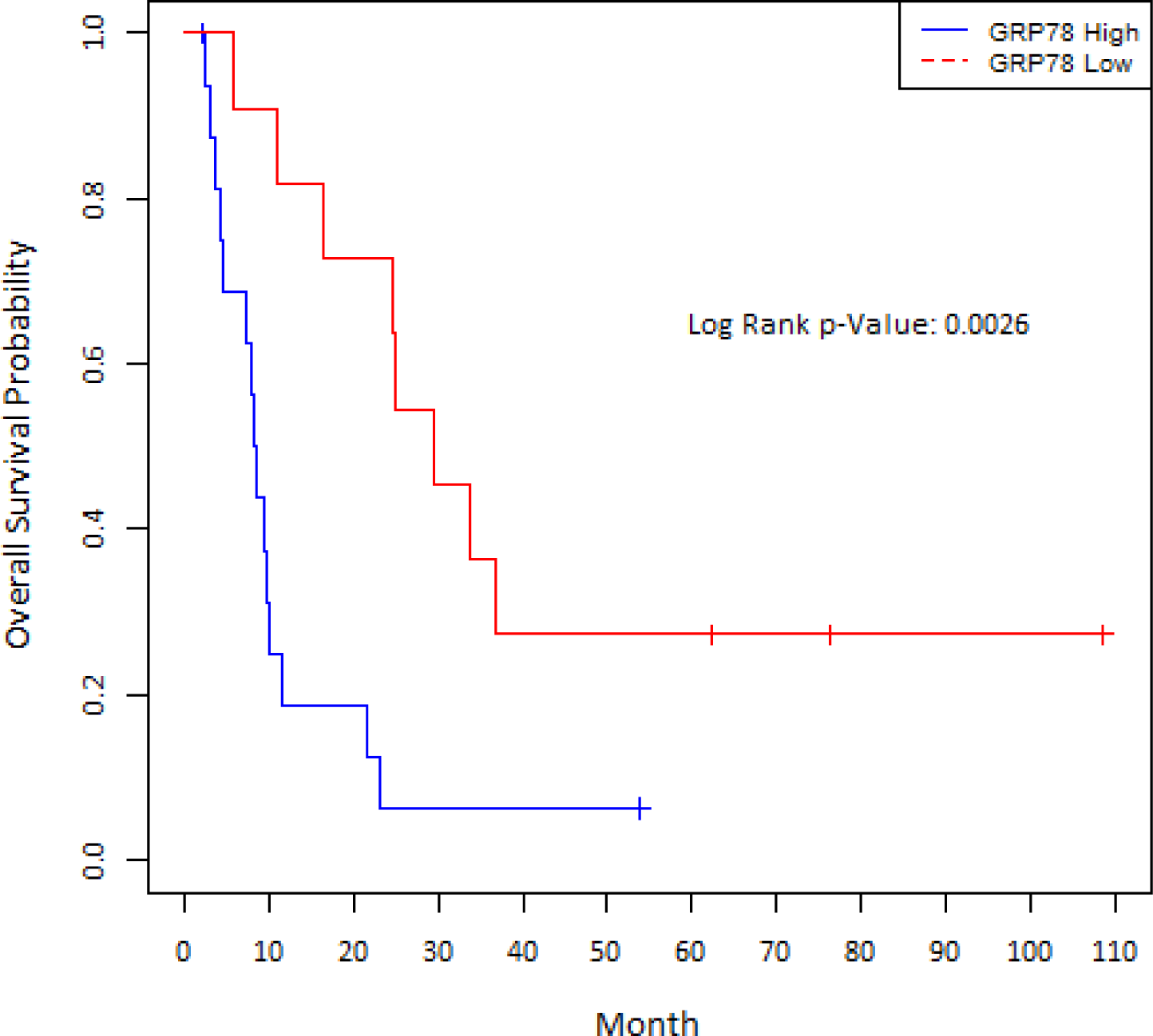
Overall Survival by GRP78 Status (high, low). P < 0.0026 of the Log-Rank Test over the strata indicates that the distributions of the overall survival time were significantly different between the high and low GRP78 patient groups.

For the five long-term surviving patients in the Kaplan-Meier tail their mean PFS was 24.9 months, and their mean OS was 73.6 months. As of March1^st^ 2024 all five long-term surviving individuals remain alive (Table 2) (Supplementary).

**Table 2:** Patient survival data for the 33 evaluable patients.

| Variable | N | Median | Mean | Minimum | Maximum |
| --- | --- | --- | --- | --- | --- |
| OS_Month | 33 | 10.0 | 21.8 | 2.1 | 108.7 |
| PFS_Month | 33 | 3.7 | 7.00 | 1.3 | 76.3 |

### Safety and tolerability

The most common AE from the combination of sildenafil, sorafenib, and valproic acid was maculopapular rash (6.4%). The three-drug combination was not associated with any grade 4 or grade 5 toxicities (Table 3).

**Table 3:** Treatment-Related Adverse Events -- Sildenafil, Sorafenib, and Valproic Acid.

| Toxicity Category | Toxicity | Number of Patients (%) |  |  |  |
| --- | --- | --- | --- | --- | --- |
|  |  | Grade1 | Grade2 | Grade3 | Total |
| Blood and lymphatic system disorders | Anemia | 1 (2.1) | 0 (0.0) | 0 (0.0) | 1 (2.1) |
| Gastrointestinal disorders | Dyspepsia | 2 (4.3) | 0 (0.0) | 0 (0.0) | 2 (4.3) |
|  | Nausea | 0 (0.0) | 1 (2.1) | 0 (0.0) | 1 (2.1) |
| General disorders and administration site conditions | Malaise | 1 (2.1) | 0 (0.0) | 0 (0.0) | 1 (2.1) |
| Immune system disorders | Allergic reaction | 0 (0.0) | 1 (2.1) | 0 (0.0) | 1 (2.1) |
| Investigations | Alanine aminotransferase increased | 0 (0.0) | 1 (2.1) | 0 (0.0) | 1 (2.1) |
|  | Aspartate aminotransferase increased | 1 (2.1) | 0 (0.0) | 0 (0.0) | 1 (2.1) |
|  | Lymphocyte count decreased | 0 (0.0) | 1 (2.1) | 0 (0.0) | 1 (2.1) |
|  | Neutrophil count decreased | 1 (2.1) | 0 (0.0) | 0 (0.0) | 1 (2.1) |
|  | White blood cell decreased | 1 (2.1) | 1 (2.1) | 0 (0.0) | 2 (4.3) |
| Metabolism and nutrition disorders | Dehydration | 1 (2.1) | 0 (0.0) | 0 (0.0) | 1 (2.1) |
|  | Hypoalbuminemia | 1 (2.1) | 0 (0.0) | 0 (0.0) | 1 (2.1) |
|  | Hypocalcemia | 2 (4.3) | 0 (0.0) | 0 (0.0) | 2 (4.3) |
| Skin and subcutaneous tissue disorders | Rash maculo-papular | 2 (4.3) | 0 (0.0) | 1 (2.1) | 3 (6.4) |

## Discussion

Our earliest pre-clinical studies with sorafenib and HDAC inhibitors determined that death receptor signaling and macroautophagy played key roles in the agents synergizing to kill tumor cells [9, 10]. This drug combination also radiosensitized tumors, prolonging animal survival [7]. Subsequently, combining sorafenib and sildenafil we demonstrated that a portion of the mechanism by which sorafenib killed tumor cells was by inhibition of PDGF receptors [23]. We then identified chaperone proteins, and in particular the chaperone GRP78, as another important target of sorafenib [17, 18]. Prolonged inhibition of GRP78 causes a persistent induction of ER stress signaling which is toxic [21, 22]. In the clinic, patients who had low GRP78 expression exhibited a significantly greater OS (p < 0.0026). Our phase 2 trial was relatively under-powered, and no significant impact on survival was associated with PDGFRα expression levels. However, had more patients been recruited to increase power, and had they shown similar profiles, it is probable that those patients with high levels of PDGFRα would also have reached significance for survival (p < 0.0728). The fact would still likely remain though that GRP78 is at least an order of magnitude more significant as a response biomarker for our drug combination than PDGFRα.

GRP78 is a multi-target chaperone that not only regulates ER stress signaling but contributes to the regulation of multiple intracellular signaling pathways [21, 22, 25–27]. GRP78 can be found in the plasma membrane, inhibition of GRP78 reduces upstream signaling into the PI3K pathway and knock down of GRP78 causes the expression of mutant KRAS proteins to decline [25, 26]. GRP78 can translocate to the nucleus and regulate transcription of the epidermal growth factor receptor (EGFR) [28]. As would be expected as a regulator of EGFR, RAS, and PI3K signaling, knock down or small molecule inhibition of GRP78 enhances tumor cell chemosensitivity [29–32]. Equally, over-expression of GRP78 stabilizes protein expression, suppresses ER stress signaling and maintains tumor cell viability. Finally, and part of the earliest studies on the protein, GRP78 also plays an essential role in the reproductive biology of all known human viruses, including SARS-CoV-2 [33, 34].

As can be envisioned, because of its pleiotropic effects, the biology linking GRP78 inhibition to tumor cell death by any drug combination is highly complex. Based on many pre-clinical studies, a significant correlation between lower GRP78 levels and prolonged survival per se is not a surprising finding, and vice versa. In our analyses, we were unfortunately unable to stratify patients based on the location of GRP78 within cells. Hence, we cannot definitively correlate whether the prolonged patient survival we observed is due to ER-localized GRP78 or plasma membrane-localized GRP78 or to nucleus-localized GRP78. Although we do not know the precise role(s) of GRP78 in reducing the efficacy of the three-drug combination, it is congruent with the concept of this chaperone facilitating plasma membrane growth factor receptor activation with greater signaling through the cytoprotective PI3K/AKT pathway.

Future studies will require many additional pre-clinical and clinical studies to fully understand our findings. Several years ago, we presented evidence that compared PDX GBM cells to selected stem-cell-like PDX GBM cells. The stem-like cells had increased their basal levels of GRP78 and drug-efflux pumps [17]. Treatment of the stem-like cells with sorafenib and sildenafil rapidly reduced GRP78 and pump levels to near those in unselected cells. Hence, the relative importance of GRP78 to OS could be due to its basal expression between patients and the number of stem cells in the glioblastoma tumor. A correlation between greater survival and elevated PDGFRα expression was approaching significance, and a new trial, with 60-100 evaluable patients, would likely show this interaction to be significant. Unlike GRP78, this finding argues that for glioblastoma cells addicted to signals from PDGFRα, the greater the expression of the receptor, the more addicted, and the more likely they are to be killed by inhibition of that receptor.

For the five surviving patients in the Kaplan-Meier tail, with a PFS of ∼25 months and an OS of ∼74 months, our drug combination played an important role in prolonging their survival. At present, we do not know the evolutionary survival mechanisms that will be present in glioblastoma cells previously treated with sorafenib, valproic acid, and sildenafil. Mouse studies using pancreatic tumor cells combining sorafenib and the HDAC inhibitor vorinostat showed activation of the EGFR family of tyrosine kinases in the surviving cells [34]. Based on likely normal tissue toxicities, it is improbable that a four-drug combination could be contemplated for recurrent glioblastoma patients, e.g., sorafenib, valproic acid, sildenafil, and the pan-EGFR inhibitor neratinib. In the future, other drug combinations similar to sorafenib, valproic acid, and sildenafil could be developed. The class III receptor tyrosine kinase inhibitor pazopanib docks with the HSP90 ATPase site and inhibits HSP90 more potently than sorafenib. Pazopanib also more potently inhibited GRP78 and HSP70 family chaperones than sorafenib [18]. Sildenafil enhanced the efficacy of both sorafenib and pazopanib to inhibit chaperone function. Pazopanib, mechanistically in a near-identical fashion to sorafenib, interacted with HDAC inhibitors to kill tumor cells [36]. However, because of overlapping normal tissue toxicities in the liver for both pazopanib and valproic acid, a combination using an HDAC inhibitor without dose-limiting liver toxicity would be required.

Another aspect of the evolutionary survival response to sorafenib, valproic acid, and sildenafil exposure is that the drug combination may be profoundly altering the epigenetic landscape in the surviving glioblastoma cells. In studies performed since completion of this trial, we discovered that drugs/drug combinations that strongly induce autophagosome formation can rapidly reduce through autophagy the expression of multiple HDAC proteins, particularly HDAC1, HDAC2, HDAC3, and HDAC6 [37–39]. Altered expression of HDACs1/2/3 can result in elevated levels of immunotherapy biomarkers, which could facilitate a long-term anti-tumor immune response. In a mouse model of triple-negative breast cancer, two weeks after cessation of neratinib and valproic acid exposure, drug-treated tumor cells still expressed less EGFR, KRAS, NRAS, HDAC1, HDAC2, HDAC3, HDAC6, and HDAC10, expressed more MHCA, than vehicle control cells. Neratinib and valproate-treated tumors had a greater infiltration of CD8+ cells [37]. Prolonged down-regulation of proteins that act to maintain cell survival such as the EGFR will increase the likelihood that the anti-tumor actions of any subsequent anti-tumor therapeutic intervention will be magnified. Whether this priming effect occurs in glioblastoma cells and whether it was responsible for increased efficacy of other modalities in this patient group will require considerable additional pre-clinical work to understand the mechanism(s).

## Data Availability

All data produced in the present study are available upon reasonable request to the authors

## Acknowledgements

This trial was supported by research funding from the NIH-NCI Cancer Center Support Grant (P30CA016059; PI: or Robert Winn, MD).

Services in support of this research were provided by the VCU Massey Comprehensive Cancer Center Biostatistics Shared Resource, supported in part with funding from NIH-NCI Cancer Center Support Grant P30-CA0016059.

The authors would like to acknowledge physicians who referred patients to the study, Zhijian Chen, MD, PhD; Alicia Zukas, MD, PhD; William Broaddus, MD,PhD; Timothy Harris, MD, PhD, Asad Khan, MD.

The authors have no conflict of interest to disclose.

## Authors’ contributions

Andrew Poklepovic: Develop and conduct study, collect and analyze data, write paper; Palak Shah: Collect and analyze data, write paper; Mary Beth Tombes: Develop and conduct study, collect and analyze data; Ellen Shrader: Develop and conduct study, collect and analyze data; Dipankar Bandyopadhyay: Develop study, analyze data, write paper; Xiaoyan Deng: Analyze data, write paper; Catherine Roberts: Analyze data, write paper; Alison Ryan: Conduct study; Daniel Hudson: Develop study and collect data; Heidi Sankala: Develop study and write paper; Maciej Kmieciak: Correlative studies; Paul Dent: Develop study, correlative studies, write paper; Mark Malkin Develop and conduct study, collect and analyze data, write paper.

## Data availability

The datasets generated during and/or analyzed during the current study are available from the corresponding author on reasonable request.

