## Supplemental data for "Phase 2 Study of Sorafenib, Valproic Acid, and Sildenafil in the Treatment of Recurrent High-Grade Glioma"

Supplementary

Note that the subject numbers used below were assigned for convenience for the purposes of this summary only and do not reflect the order of these or any other subjects’ clinical trial participation.

Additional details regarding the disease phenotypes may be obtained by contacting the corresponding author.

Subject 1 was initially diagnosed with oligodendroglioma, WHO grade 2 to 3, in 1998 while in his 30’s. He underwent resection at that time and again in 2012 for a recurrence, which was consistent with high grade glioma, anaplastic oligodendroglioma, WHO grade 3. Following progression after 7 cycles of temozolomide, he completed 4 cycles of procarbazine, lomustine, and vincristine, then discontinued due to side effects. In 2014 Subject 1 was noted to have progression and was then enrolled on this clinical trial; he remained on trial until 2016 (received 19 cycles) when tumor progression was noted. GRP78 staining showed a score of 0. Post-trial, Subject 1 had 3 months of radiation therapy in 2016 and 6 cycles of bevacizumab from 2016 to 2017. Incidentally in 2022, a right tentorial meningioma was found and progressed 6 months later; radiation to the meningioma was completed in 2023. Currently, in his 50’s, Subject 1 is on surveillance. Given that it has been 7 years since his second progression, it is unclear if his excellent survival is due to this three-drug regimen, due to the low GRP78 score, and/or as an outlier.

Subject 2 was diagnosed with right temporoparietal glioblastoma WHO grade 4, 2016, in his 50’s. He underwent resection in 2016, followed by concurrent radiation and temozolomide and then adjuvant temozolomide for 4 months, with tumor progression. A surgical biopsy with laser ablation was performed in 2017 revealing recurrent glioblastoma; Subject 2 was then enrolled on this clinical trial in 2017 and continued until 2023. GRP78 staining showed a score of 1. Participation was stopped after 7 cycles due to evidence of disease progression on radiographic imaging. However, later in 2023, Subject 2 had a biopsy at another institution that did not show any evidence of malignancy. He has since remained off treatment with serial imaging showing radiological stability. The laser ablation prior to trial participation could have led to his survival, but this three-drug combination or the low GRP78 score possibly could have contributed to survival as well.

Subject 3 was diagnosed with left occipital glioblastoma, WHO grade 4 in 2017, in her 50’s with resection that year. She completed radiation therapy in 2017 but had evidence of tumor progression and received adjuvant temozolomide for one cycle with progression. Subject 3 was enrolled on this clinical trial in 2018 and continued for 6 months; receiving 6 cycles, when radiographic progression was noted on MRI. GRP78 staining score was 1. She had a dose reduction of sorafenib to 200mg twice a day due to adverse events of painful foot blisters during the trial. She then received bevacizumab and lomustine for 10 months in 2018 to 2019. Since then, she has had clinical and radiographical stability with interval MRI imaging. The progression noted on the scans in 2018 could have been radiation necrosis.

Subject 4 was diagnosed with anaplastic astrocytoma, WHO grade 3, in 2008 as a teenager. He had surgical resection in 2008 along with concurrent radiation and temozolomide, followed by 13 cycles of temozolomide, ending in 2010. He had tumor progression in 2016, in his 20’s and was treated with adjuvant temozolomide from 2016 to 2017. Subject 4 had tumor progression in 2018 and was enrolled on this clinical trial from 2018 to 2020; participation was stopped due to slow disease progression seen on MRI, after 3 cycles. The GRP78 status is unknown. He had a surgical biopsy in 2020 showing recurrent anaplastic astrocytoma, Subject 4 had surgical resection in 2020. He was then enrolled on a phase 2 clinical trial of nivolumab for IDH mutated glioma with hypermutator phenotype from 2021 to 2022; participation was stopped due to disease progression. He had re-irradiation in 2022. A repeat biopsy later that year showed recurrent anaplastic astrocytoma, grade 3, IDH mutant. Subject 4 was then started on bevacizumab and lomustine beginning in 2023 and continuing currently. Multiple factors such as young age and IDH mutation status favor positive prognosis.

Subject 5 was diagnosed with WHO grade 3 oligodendroglioma in 2007, in his 50’s. He had multiple resections. Subsequent biopsies were as follows: later in 2007 (WHO grade 2 oligodendroglioma; 2013 WHO grade 2 oligodendroglioma; 2017 WHO grade 3 oligodendroglioma). After concurrent radiation and temozolomide for 2 months in 2018, followed by adjuvant temozolomide for 12 cycles in 2018 to 2019, Subject 5 was enrolled on this clinical trial in 2019, in his 60’s, for 2 cycles. GRP78 staining score was 1. Participation was stopped due to concern for tumor growth, but subsequent serial imaging showed evidence of radiation effect rather than tumor progression, and Subject 5 is currently on surveillance with clinical and radiographical stability.
